# Changing indications for intravitreal Anti-Vascular Endothelial Growth Factor Injections at the University College Hospital, Ibadan, sub-Saharan Africa

**DOI:** 10.1101/2020.06.18.20135129

**Authors:** A.T Adewole, T.S Oluleye, Y.O Babalola, O Majekodunmi, M Ijaduola

## Abstract

**Aim:** To compare the current indications for intravitreal anti-vascular endothelial growth factor (anti-VEGF) therapy, to make recommendations for planning of services.

**Methodology:** The medical records of 172 patients who had intravitreal anti-VEGF injections from January 2016 to December 2019 were retrieved. Socio-demographic and clinical data were extracted, analysed, and compared with data from the previously published audit report covering 2010 to 2012.

**Results:** Three hundred and thirty injections were given to 182 eyes in this cohort of patients. The mean age was 61.1 ± 16.3 years (range <1-90 years), with a male to female ratio of 1.1:1. Retinal vein occlusion, 64 (35%) remained the most common indication for anti-VEGF injections in the eyes treated. This was followed by choroidal neovascular membrane/ Wet age-related macular degeneration which accounted for 42 (23%) as reported previously. However, cases of proliferative diabetic retinopathy/ diabetic maculopathy needing anti-VEGF were noticed to have almost doubled from about 8% (10) in the previous study to 15% (27) in the present study. In addition, idiopathic polypoidal choroidal vasculopathy, 18 (10%) ranked above proliferative sickle cell retinopathy in the present study. Retinopathy of prematurity, neovascular glaucoma, retinal artery macro-aneurysm and myopic choroidal neovascular membrane were the new emerging indications.

**Conclusion:** There is an expanding indication for anti-VEGF in the management of retinal vascular diseases in our health facility and adequate measures should be put in place for early diagnosis and management. Patients should be counselled on the availability of this treatment option.

## Introduction

The use of anti-VEGF therapy is an effective therapy in many clinical trials for choroidal neovascular age-related macular degeneration (wet AMD), diabetic macular oedema (DME), macular oedema due to retinal vein occlusion (RVO), myopic choroidal neovascularisation (myopic CNV) and other retinal diseases.^1^ As a result, anti-VEGF has become the standard treatment for various retinal vascular diseases, either as first or second-line option^1,2,3,4^. The aim of this clinical audit is to compare the current indications for intravitreal anti-VEGF treatment in University College Hospital, Ibadan with the previous audit done 5 years ago for the purpose of making recommendations for improved eye-care services.^5^

## Methodology

This retrospective study included 182 eyes of 172 patients who had intravitreal anti-VEGF treatment in the Vitreo-retinal unit of the hospital between 2016 and 2019. This study adhered to the tenets of the Declaration of Helsinki; informed consent was obtained from the patients before the injections under aseptic condition in an operating theatre. Periocular skin and eyelids were cleaned with 10% povidone-iodine while 5% povidone-iodine drop was instilled into the conjunctiva sac after topical tetracaine. The injections were given through the pars plana (4mm for phakics and 3.5mm for pseudophakics; 1.5mm for neonates). A dose of 1.25 mg/0.05ml for adults and 0.625mg for neonates of bevacizumab (Avastin, Roche, Basel, Switzerland) was used while 0.5 mg/0.05ml for adults and 0.25mg for neonates for ranibizumab (Lucentis, Novartis, Basel, Switzerland) was used. All patients consented to the Off-label use of bevacizumab (Avastin) and were given topical moxifloxacin qid for 1 week after each injection.

Socio-demographic and clinical data were extracted from the patients’ medical records and compared with data from the previously published audit report on indications for anti-VEGF injections.^6^ Data were analysed using the Statistical Package for Social Sciences IBM (SPSS-IBM), version 24 (SPSS Inc., Chicago, Illinois, USA), and reported as frequency distributions and percentages.

## Results

Three hundred and thirty injections were given to 182 eyes to this cohort of patients. The mean age was 61.1 **±** 16.3 years (range <1-90 years), with a male to female ratio of 1.1:1. Majority of patients that took injections were older than 60 years in both reviews (Figure 1). Retinal vein occlusion (35%) remained the most common indication for anti-VEGF injections followed by choroidal neovascular membrane/ Wet age-related macular degeneration (23%) but in higher proportion when compared to the previous study. Proliferative diabetic retinopathy and diabetic macular oedema (15 %) and idiopathic polypoidal choroidal vasculopathy (10%) also ranked above sickle cell retinopathy (3.3%). Retinopathy of prematurity, neovascular glaucoma and retinal artery macro-aneurysm were the new emerging indications observed in this study (Table 1).

**Table 1:** Comparison of indications for anti-Vascular Endothelial Growth Factor.

| Indications |  | 2016-2019 (Year) |  | 2010-2012 (Year) |  |
| --- | --- | --- | --- | --- | --- |
|  |  | Frequency(n=182) | Percentage (%) # | Frequency(n=134) | Percentage (%) |
| Retinal Vein Occlusion +CME | BRVO | 34 | 7.7 | 26 | 19.4 |
|  | CRVO | 16 | 18.7 |  |  |
|  | HCRVO | 14 | 8.9 |  |  |
| Wet Age-related macular degeneration |  | 42 | 23.0 | 23 | 17.1 |
| IPCV |  | 18 | 9.9 | 8 | 6 |
| PDR/DME |  | 27* | 14.8 | 10 | 7.5 |
| CME (Non-diabetic related) |  | 9 | 4.9 | - | - |
| Proliferative SCR |  | 6 | 3.3 | 22 | 16.4 |
| RAM |  | 4 | 2.2 | 6 | 4.5 |
| ROP |  | 3 | 1.7 | - | - |
| NVG |  | 2 | 1.0 | 7 | 5.2 |
| Idiopathic CNVM |  | - | - | 7 | 5.2 |
| Others |  | 7* | 3.9 | 2 | 1.5 |

CRVO= central retinal vein occlusion; HRVO= hemi-retinal vein occlusion; BRVO= branch Retinal Vein Occlusion; IPCV= idiopathic Polypoidal Choroidal Vasculopathy; PDR= proliferative diabetic maculopathy; DME= diabetic macular oedema; CME= cystoid macular oedema; SCR= sickle cell Retinopathy; RAM= retinal arterial macroaneurysm; ROP= retinopathy of prematurity; NVG= neovascular glaucoma; CNVM= choroidal neovascular membrane
\* Others: Vitreous haemorrhage in POAG, Presumed Toxoplasmosis, Choroidal melanoma, Intraretinal mass (unspecified), Haemorrhagic macular detachment, Specific uveitis, Myopic CNVM

**Figure 1:**
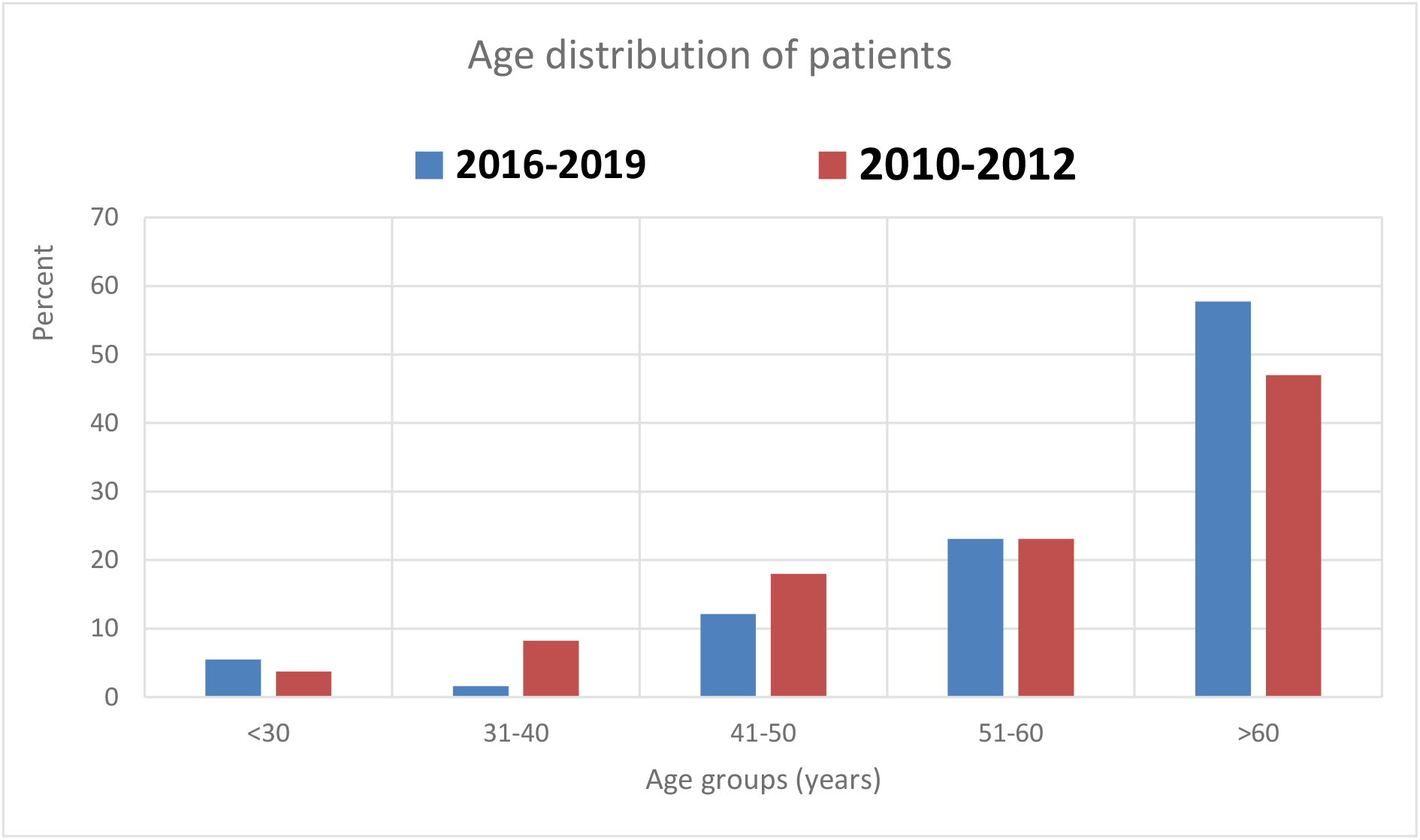
Age distribution of patients.

## Discussion

Retinal venous occlusions remained the most common indication for anti-VEGF injections in our centre. This is similar to a study Fiebai et al,^7^ but at variance with the report from Port-Harcourt and from other centres in Nigeria where diabetic retinopathy/ diabetic maculopathy was reported as the most common indication.^8,9,10,11^ This disparity may be from the peculiarity of our centre (University College Hospital, Ibadan) being a larger centre that receives more referral from many states in the country. Surprisingly, the proportion of patients treated for diabetic retinopathy had doubled over the 5 years when compared to the last audit. Furthermore, the increasing prevalence of diabetes mellitus and its accompanying eye diseases could have accounted for the increase in these cases.^12,13^ This could be due to increased partnership with the Endocrinology unit in the hospital. This partnership has resulted in the expansion of ophthalmic screening services and direct referrals to the eye clinic at the time of diagnosis in the endocrinology clinic. Also, a memorandum of understanding with the hospital management enabled the members of the Diabetic Association to get a 50% price reduction for fundus photo screening services at the eye unit of the hospital.

Choroidal neovascular membrane (CNVM) mainly from age-related macular degeneration and other aetiology like myopic CNVM was also observed as a major indication for anti-VEGF treatment. This may have been supported by the fact that anti-VEGF therapy remains a mainstay of treatment. ^3,14,15^ Similarly, advances in the medical field that has led to the increased population of the elderly who may have accounted for the cases of wet ARMD.^16^ On the other hand, the proportion of patients with proliferative sickle cell retinopathy (SCR) needing intravitreal anti-VEGF has significantly reduced in our centre (from 16% to 3% over 5 years). This may have been largely due to the availability of retinal laser services for less complicated cases and use of vitrectomy for non-clearing vitreous haemorrhage from proliferative SCR. Access of patients to proper counselling may be responsible for improved health-seeking behaviour.

We noticed the increasing proportion of cases of Idiopathic polypoidal choroidal vasculopathy (IPCV) managed with anti-VEGF in this review. This is similar to the report by Oluleye et al,^17^ in their case series where IPCV was found to be common in sub-Saharan but treated with vitrectomy or referred abroad. This increased number treated with anti-VEGF may be due to earlier presentation and improved diagnosis with the availability of optical coherence tomography scan in the hospital. Those cases previously diagnosed as the idiopathic choroidal neovascular membrane in 2014 were cases of IPCV.

Retinopathy of prematurity (ROP), neovascular glaucoma and retinal artery macro-aneurysm were the new emerging indications in this review. Treatment of ROP with anti-VEGF stems from a collaborative initiative by the retinal unit, paediatric ophthalmology, and the paediatric units in the hospital for a sustainable ROP screening service. Hence, anti-VEGF has been added to the armamentarium of this childhood blinding disease as a preferred practice pattern as corroborated by previous studies.^18–22^

Myopic CNVM was also noted to be an emerging indication that would likely be on the increase as the global myopia pandemic wave runs through sub-Saharan Africa. ^23–26^ This trend has tremendous implications for planning services, including managing and preventing myopia-related ocular complications and vision loss among almost 1 billion people with high myopia in which anti-VEGF will probably be a major player.^26^

## Conclusion

There is an expanding role of anti-VEGF in the management of retinal vascular diseases in Ibadan. Appropriate planning should be put in place to accommodate the increasing cases for diabetic retinopathy, retinopathy of prematurity and other emerging diseases. Patients should be counselled on the availability of this treatment option.

## Data Availability

Data are available in SPSS file on request through the corresponding author and approval by the Hospital management.

## Footnotes

### Limitations

The study being a retrospective study, may be limited by accurate data retrieval from case records.

### Conflicts of interest

Authors report none.

### Acknowledgements/Financial Support

There was no external financial support for this project.

### Contributions

We thank Mr. Iwa of the records department for the excellent work done in retrieving the case notes. Appreciation goes to Dr. Ibiyemi (Ophthalmology) and Mr. Seun Ayangbesan (Research assistant) who helped in data extraction. TO, YOB, OM, MI, and AA contributed to the design of the study. AA analysed the data and prepared the initial manuscript. All authors reviewed, proofread, and approved the final manuscript and references.

### Patient consent

Obtained

## Notes

### Competing Interest Statement

The authors have declared no competing interest.

### Funding Statement

No external funding was available for this review

### Author Declarations

University of Ibadan/University College Hospital Ethical Review Board

